# A Dialectical Behavior Therapy – Skills Training Intervention for Cigarette Smoking by Patients with Cancer: Qualitative Descriptive Study of Intervention Components Prior to Feasibility Trial

**DOI:** 10.1101/2025.08.19.25334013

**Authors:** Marcia H. McCall, Charlotte T. Boyd, Nicole D. Kerr, Sherona R. Sirkisoon, Ellen A. Nicola, Erica Fox, Lindsay K. Seigenthaler, Stephanie S. Daniel, Erin L. Sutfin

## Abstract

**Background:** Despite evidence that quitting smoking improves cancer treatment and outcomes, about one in eight patients with cancer smokes cigarettes. Barriers to quitting include difficulty managing cancer-related emotions and distress without smoking and low self-efficacy for quitting. Patients who continue to smoke may benefit from targeted therapies beyond motivational and cognitive approaches commonly used in standard-of-care smoking cessation treatment. Previous studies using Dialectical Behavior Therapy – Skills Training (DBT-ST) successfully improved coping skills and outcomes for patients with substance use disorders and patients with breast cancer, but DBT-ST has yet to be applied to smoking among cancer populations. We developed an eight-session virtual group DBT-ST intervention to teach mindfulness, emotion regulation, and distress tolerance coping skills to patients with cancer who continue to smoke.

**Primary Aim:** To improve our intervention before feasibility testing in a clinical trial, we solicited critiques and commentary on key intervention components from patients with cancer and smoking histories and their multidisciplinary clinical team.

**Methods:** Within a pragmatist paradigm, we used qualitative description to conduct semi-structured qualitative interviews with two groups: patients with cancer who smoked or had quit (*n*=8) and oncology clinicians (physicians, psychotherapists, and nurses *n*=9). The interviews were analyzed using codebook thematic analysis.

**Results:** Patients and clinicians supported intervention components for smoking cessation, particularly its group format and teaching DBT-ST skills for being present, managing emotions, and handling distress. Participants noted perceived benefits including improved quality of life, decreased anxiety and depression, and acquisition of universal skills for managing life’s challenges. Other components, such as a virtual setting and the inclusion of patients not ready to quit, received mixed reviews.

**Conclusions:** Results indicate early enthusiasm for our DBT-ST intervention for patients with cancer who smoke cigarettes, supporting progression to a feasibility study following specified modifications to intervention components.

**Highlights:** Continued smoking after cancer diagnosis is associated with poorer outcomes.

We designed an original program to reduce reliance on smoking as a coping strategy.

The program teaches stress management, emotion regulation, and mindfulness skills.

Patients and oncology clinicians provided feedback on key proposed intervention components.

Feedback informed targeted intervention refinements for future feasibility testing.

## 1. Introduction

Patients who continue smoking cigarettes after their cancer diagnosis are at increased risk of death, failed treatment, treatment-related complications, and second primary cancers compared to patients who stop smoking (Caini et al., 2022; Cinciripini et al., 2024; National Center for Chronic Disease et al., 2014; Tohmasi et al., 2025; United States Public Health Service Office of the Surgeon et al., 2020). In contrast, smoking cessation after diagnosis is associated with several benefits, including improved survival, better surgical outcomes, lower risk of recurrence or second primary cancers, reduced fatigue, and increased psychosocial well-being (Caini et al., 2022; Chellappan, 2022; Cinciripini et al., 2024; Lucchiari et al., 2016; Mohebbi et al., 2025). Although a cancer diagnosis can be a catalyst for cessation (Puleo et al., 2022), approximately 12.7% of patients with cancer continue to smoke cigarettes (Salloum et al., 2019).

Clinical practice guidelines recommend a combination of behavioral counseling (such as motivational or cognitive therapy) and pharmacotherapy for smoking cessation in oncology settings (National Comprehensive Cancer Network, 2022). These interventions help patients manage withdrawal symptoms, identify and plan for situations that trigger cravings, align motivation to quit with personal values and goals, and build confidence in their ability to quit (National Comprehensive Cancer Network, 2022). Although this approach is effective for many patients with cancer, particularly when delivered with greater intensity, such as six to eight counseling sessions plus medication (Cinciripini et al., 2019; Cinciripini et al., 2024), a subset of patients with cancer struggle to quit smoking, including many receiving evidence-based treatment (Duffy et al., 2012; Sheeran et al., 2019; U.S. National Cancer Institute, 2022). Factors associated with continued smoking include psychological distress, low cessation self-efficacy, cancer- and smoking-related stigma, and difficulty tolerating nicotine withdrawal symptoms (Duffy et al., 2012; Sheeran et al., 2019; U.S. National Cancer Institute, 2022). Furthermore, cancer diagnosis and treatment can induce or worsen distress, anxiety, depression, hopelessness, and interpersonal conflict (Duffy et al., 2012; Lee et al., 2021; Lucchiari et al., 2016; Wells et al., 2017), conditions that are associated with fewer quit attempts and higher rates of relapse among patients with cancer (Duffy et al., 2012; Lee et al., 2021; Schlam et al., 2020). For these patients, cigarette smoking can serve as a primary means of coping with negative experiences (Duffy et al., 2012; Wells et al., 2017). Consequently, developing interventions that address these challenges has been identified as a national priority for improving smoking cessation outcomes in oncology populations (U.S. National Cancer Institute, 2022).

Existing motivational and cognitive approaches for smoking cessation may not directly address difficulties tolerating distress or regulating emotions. Inability to handle stress and regulate emotions are transdiagnostic processes associated with smoking persistence in the general population (Carmody et al., 2007; Leventhal & Zvolensky, 2015; Schlam et al., 2020; Veilleux, 2019). One potential mechanism underlying continued smoking among patients with cancer is a lack of skills for managing stress and negative emotions instead of relying on smoking to cope. Interventions that teach and reinforce these skills may provide alternative coping strategies, improve cessation self-efficacy, and improve smoking cessation outcomes.

### 1.1. Dialectical Behavior Therapy

A counseling approach to address these challenges is Dialectical Behavior Therapy (DBT), a mindfulness-based psychotherapy designed to help individuals develop more effective ways of responding to distress. The term *dialectical* refers to the synthesis of opposites. In DBT, this principle emphasizes balancing acceptance of oneself and one’s experiences with efforts to change emotions, behaviors, and interpersonal relationships (Dimeff & Linehan, 2008). Through skills training, DBT teaches four core competencies: mindfulness, distress tolerance, emotion regulation, and interpersonal effectiveness. Central to DBT is the goal of helping people construct a “life worth living” by developing adaptive coping strategies and increasing engagement in valued activities (Linehan, 2015). Moreover, DBT has demonstrated effectiveness across populations from varied cultural backgrounds (Cogwell Anderson et al., 2013; Harned et al., 2022; Linehan, 2015).

Because comprehensive DBT typically requires at least one year of treatment, researchers have adapted its core component, DBT Skills Training (DBT-ST), into brief and more feasible interventions. In the only known study of DBT-ST as a brief intervention specifically targeting cigarette smoking, a small trial among women receiving treatment at a methadone clinic found the intervention to be feasible and associated with reductions in cigarette use by 83% (Cooperman et al., 2019). Interpretation of these findings is limited by the small sample size (*n*=7), inclusion of women only, and lack of a control group.

Evidence from studies of populations with characteristics relevant to patients with cancer who smoke further supports the potential utility of DBT-ST. Among individuals with substance use problems and patients with breast cancer experiencing psychological distress, DBT-ST interventions have been associated with reductions in distress, anxiety, and depression, substance use, and cravings, as well as improvements in emotion regulation, distress tolerance, mindfulness skills, and cessation self-efficacy (Author1 et al., 2025; Onken et al., 2014). To our knowledge, however, DBT-ST has not been adapted or evaluated as a tobacco treatment intervention for patients with cancer.

### 1.2. DBT-ST Intervention Development

Given the need for effective for cigarette smoking cessation interventions tailored to patients with cancer, we developed a DBT-ST intervention for this population using the NIH Stage Model for Behavioral Intervention Development as a guiding framework. The NIH Stage Model describes a systematic process for developing and evaluating behavioral interventions, progressing from intervention design and refinement to efficacy and effectiveness testing (Onken et al., 2014). Stage 1A focuses on 1) identifying promising scientific findings to support intervention development, 2) designing the intervention, and 3) refining or modifying the intervention for real-world settings (Onken et al., 2014).

To identify promising scientific findings, we conducted a scoping review of DBT-ST as a brief intervention for difficult-to-treat populations: people with substance use disorders and people in severe psychological distress, including patients with cancer (Author1 et al., 2025). We found that DBT-ST demonstrated preliminary efficacy as a brief intervention for these populations (Author1 et al., 2025). To help patients with cancer gain self-efficacy for quitting smoking and preventing relapses, we designed a novel eight-session DBT-ST intervention specific to the psychological experiences and smoking behaviors of this population, teaching skills in mindfulness, distress tolerance, and emotion regulation (see **Table 1** for the original DBT-ST intervention). Our intervention aims to help patients who smoke daily and are both ready and not ready to quit smoking to build new coping skills and self-efficacy for quitting.

Patients are encouraged to reduce or quit smoking and to use pharmacotherapy to support quitting. During design, we collaborated with a multidisciplinary advisory board of oncology and family medicine physicians, nurses, behavioral health clinicians, and a patient with cancer who had quit using tobacco. The board met six times to provide intervention feedback and revision.

### 1.3. Study Goals and Research Questions

Prior to evaluating our intervention in a single-arm feasibility clinical trial, we looked to improve key intervention components by identifying potential barriers to and facilitators of success as a smoking cessation treatment program for the cancer population. To achieve this primary goal, we conducted a qualitative study involving patients with cancer who smoked cigarettes or had quit and oncology clinicians. Our secondary goals were 1) to obtain feedback on patient-reported assessments planned for the feasibility trial to evaluate tobacco use, psychological symptoms, and skills use and 2) to gather suggestions for trial recruitment and retention. Our research questions regarding intervention components were the following:

1. Primary: Is the DBT-ST brief intervention content appropriate, useful, and helpful for patients with cancer who smoke cigarettes? Are the intervention setting, format, and number and length of sessions appropriate for the patient population and sufficient for the content?
2. Secondary: Are the planned patient-reported assessments relevant to patients with cancer and appropriate for use in future clinical trials of the intervention? What recruitment and retention activities are most likely to engage and retain patients in trials of the intervention?

This study completes Stage 1A of the NIH Stage Model to refine our DBT-ST intervention for cigarette smoking by patients with cancer.

### 1.4. Study Theoretical Paradigm

Our theoretical paradigm was pragmatism as described by Morgan (Morgan, 2007, 2014). Pragmatism emphasizes understanding human experience in ways that inform practical decision-making and action, proposing that individuals reflect on their beliefs to guide actions and, in turn, reflect on the outcomes of those actions to refine their beliefs. Within this framework, emotions are viewed as important links between beliefs and actions. This perspective is particularly relevant to smoking cessation among patients with cancer, as both smoking and cancer carry profound personal and social meaning and often evoke challenging emotions that can influence health behavior.

Accordingly, participant sampling prioritized patients and clinicians with direct experience relevant to smoking cessation in cancer care. Interview guides focused on practical questions regarding intervention content, delivery, assessments, recruitment, and engagement, while also exploring how emotions, beliefs, and experiences shaped smoking behavior and cessation efforts. Data analysis emphasized participants’ descriptions of experiences, barriers, facilitators, emotional challenges, and recommendations rather than generation of explanatory theory. Consistent with the pragmatic objective of producing actionable knowledge, participant perspectives were used to refine the proposed DBT-ST intervention, ensuring that findings directly informed intervention development and future clinical research.

## 2. Methods

### 2.1. Qualitative Methodology

Using the qualitative description (QD) method as explained by Sandelowski (Sandelowski, 2000, 2010), we solicited critiques and commentary on components of our proposed DBT-ST intervention from patients with cancer who smoked or had quit and from their clinicians. QD uses cataloging, describing, and summarizing data in the “everyday language” of participants as entry points for future study. The method prioritizes natural and direct communication from participants and observed rather than covert themes or interpretations. The combination of QD and pragmatism was selected because the primary aim of this study was to generate recommendations for intervention component refinement and future feasibility testing rather than develop theory. QD aligns well with the pragmatism paradigm’s prioritization of human experience, as well as with our goal to generate findings that accurately reflect participant views and were directly relevant to intervention development and future clinical practice.

### 2.2. Study Design

This study used virtual semi-structured individual interviews with patients with cancer who smoked cigarettes or had quit and with oncology clinicians. COREQ/SRQR reporting guidelines (O’Brien et al., 2014; Tong et al., 2007) were followed (see Supplementary Materials **S1**. COREQ Checklist and **S2**. SRQR Checklist). This study’s design and its analysis were not pre-registered.

### 2.3. Ethical Approval and Participant Consent

The Institutional Review Board (IRB) of record provided ethical approval (IRB00109328). Potential participants reviewed the IRB approved study information sheet and verbally consented to study participation.

### 2.4. Setting

The setting for this qualitative study was a large comprehensive cancer center in the southeastern United States.

### 2.5. Participants

We recruited participants from the patient and clinician populations of the cancer center. We sought a sample that was racially and ethnically representative of the cancer center’s population of patients who smoked, and a clinician sample with diverse professional backgrounds. This purposive sampling process was consistent with guidance for using QD methods in health science research (Colorafi & Evans, 2016).

#### Patients

Potential patient participants were recruited by medical and radiation oncologists, psychosocial oncology therapists, and cancer center study coordinators and referred to the research team for screening. Included were English-speaking adult patients aged 18 years or older with cancer, active or in remission, who were currently smoking or had quit. Participants who previously smoked were included as evidence shows this population can elicit feedback on the most impactful drivers for quitting smoking (Hughes, 2003; Murphy-Hoefer et al., 2018; Wang et al., 2014). We excluded patients with cognitive impairment or altered cognition due to psychiatric illness or illicit psychoactive drug use. Our cancer center population of patients who smoke is approximately 22% non-Hispanic Black/African American (hereafter, Black) and 76% non-Hispanic White/Caucasian (hereafter, White). Patients satisfying screening criteria were contacted either in person on their oncology visit or by phone to learn about the study and complete the consent and enrollment process. Patient participants received $10 gift cards for their participation.

#### Clinicians

We contacted clinicians from diverse specialties and professions who were known to us or suggested by other clinicians. All clinicians of the cancer center were eligible to participate, including oncology physicians, advanced practice providers, nurses, and psychotherapists. Clinicians who responded affirmatively received information about the study and were consented via phone. Clinician participants were not compensated for taking part in this study.

### 2.6. Interviews and Interviewers

Consistent with both pragmatism and QD, we conducted semi-structured interviews intended to solicit direct understanding of patient experiences with cancer and smoking, as well as how the proposed intervention components might inform and improve their experiences (Colorafi & Evans, 2016; Morgan, 2014). Interview guides created structure and were developed through multiple iterations by our independent in-house qualitative research service, Qualitative and Patient-Reported Outcomes (Q-PRO), in collaboration with two members of the research team, the Principal Investigator (PI, a doctorate-level Licensed Clinical Mental Health Counselor [LCMHC]) and a master’s-level family medicine health educator (Certified Health Education Specialist). The guides were not pilot tested prior to data collection; however, no revisions were necessary during the interview process (see Supplementary Materials for **S3**. Patient Interview Guide and **S4**. Clinician Interview Guide).

Four team members, including the health educator and three cancer center LCMHCs, conducted interviews. Interviewers were not formally training in DBT-ST, nor did they conduct DBT-ST treatment with their patients, but they gained rudimentary DBT-ST knowledge from helping develop the intervention. Their acquaintance with DBT-ST allowed them to answer questions, clarify intervention components, and offer relevant context for application of DBT skills to the target population. The interviewers’ professional clinical training provided essential interview skills in reflective listening, empathetic discussion, and skilled communication. They followed up participant responses with clarifying or probing questions to fully elucidate participant experiences, per QD methods (Sandelowski, 2000).

#### Interview Content

Participants were informed sufficiently about the intervention components to provide knowledgeable feedback and were encouraged to share their honest opinions regarding the components in order to best prepare it to be feasible for patients undergoing treatment for cancer and acceptable or worth pursuing for someone trying to reduce or quit smoking (Kallio et al., 2016). Questions about interview structure focused on the group format, virtual setting, and session length and whether to include patients at all stages of treatment and readiness to quit smoking. Questions about interview content sought feedback on DBT-ST skills. To describe the skills, interviewers read excerpts from the intervention manual. These included descriptions of mindfulness, emotion regulation, and distress tolerance and how these coping skills could help patients deal with the emotions and stress of having cancer rather than relying on smoking to cope. Interviewers asked patient participants to reflect on how these components related to their cancer and smoking, and whether they perceived the skills as useful and feasible to learn.

Interviewers described mindfulness skills as allowing a patient with cancer to “press the pause button” when upset and regain the ability to make healthy behavior choices. They presented emotion regulation skills as helping patients during emotionally intense moments and restoring healthy functioning. Distress tolerance skills could help patients make stressful or painful situations more tolerable, enabling them to refrain from behaviors that could make situations worse. Recruitment and engagement questions focused on barriers and facilitators to patients with cancer who smoke enrolling in, engaging with, and completing the intervention program, and how to best recruit this patient population to clinical trials.

A battery of validated, open-access psychometric assessments was selected for potential use in the future feasibility study. Measures were chosen from those identified in the scoping review (see **Table 2** for the assessments, associated items, and estimated completion times). Patient participants completed the assessment battery, whereas clinician participants reviewed the assessments. The battery measured tobacco use, emotion regulation, distress tolerance, depression, anxiety, stress, and adjustment to cancer. It included the Cancer Patient Tobacco Use Questionnaire (C-TUQ; 22 items) (Land et al., 2016), the core C-TUQ smoking-status questions intended for repeated assessment of recent tobacco use, the Difficulties in Emotion Regulation Scale (DERS; 36 items) (Burton et al., 2022; Hallion et al., 2018), the Distress Tolerance Scale (DTS; 15 items) (Simons & Gaher, 2005), the Mental Adjustment to Cancer Scale (MAC; 40 items) (Watson et al., 1988), and the Depression Anxiety Stress Scales (DASS-21; 21 items) (Crawford & Henry, 2003; Lovibond, 1995). Completion of the full battery required approximately 24 to 43 minutes. Participants evaluated the measures’ relevance, comprehensibility, acceptability, and anticipated respondent burden; they were not asked to evaluate the measures’ psychometric properties or suitability as trial outcome measures.

### 2.7. Reflexivity

Factors relevant to interviewer reflexivity included interviewers’ clinical backgrounds in health education and psychotherapy, familiarity with DBT-ST, involvement of several interviewers in intervention development, and shared positionality as White women in early- to mid-career stages. Interviewers entered the study with the assumption that DBT skills could support smoking cessation among patients with cancer and recognized that their perspectives and participants’ awareness of their affiliation with the intervention development team could influence interview responses and interpretation. To mitigate these influences, interviewers completed two hours of qualitative interviewing training with a Q-PRO qualitative research expert before data collection. Training addressed common interviewing pitfalls, including leading questions and completing participants’ thoughts, and emphasized neutral, non-leading language, appropriate use of pauses, and prompts that encouraged elaboration. Interviewers were instructed to elicit both supportive and critical feedback, probe concerns and barriers, and invite suggestions for modifying components perceived as unacceptable, burdensome, or unlikely to be effective. To minimize the influence of pre-existing relationships, interviewers did not interview patients known to them or clinicians with whom they had working relationships; all but two clinician interviews were conducted by the family medicine health educator, who had no prior interactions with clinician participants. Clinicians completed interviews in clinic or home offices, and patients completed interviews in their homes; to the interviewers’ knowledge, no other individuals were present. In addition, an external qualitative researcher who had not participated in intervention development contributed to coding and theme development and independently examined the developing interpretations, including alternative explanations and disconfirming cases.

### 2.8. Data Collection and Participant Privacy

Individual semi-structured interviews were conducted by telephone or videoconference and audio-recorded by one of four interviewers, who prepared field notes immediately after each interview. Participants were encouraged to complete interviews in a private location of their choosing. A professional transcription service transcribed the recordings verbatim, and a study team member verified transcript accuracy against the corresponding audio recordings. Participants were informed that they might be contacted to clarify transcript content or provide additional information if needed; however, no such follow-up was required. Participant validation (member checking) was not conducted because it was not congruent with the study’s pragmatism and qualitative descriptive approaches. Methodological rigor and trustworthiness were instead supported through an audit trail and a robust data analysis plan, which also helped mitigate the influence of individual researcher perspectives. Audio recordings and transcripts were stored on secure, institutionally managed servers accessible only to authorized study personnel. Transcripts were de-identified before analysis, and quotations selected for dissemination were reviewed to remove potentially identifying details while preserving their substantive meaning.

### 2.9. Data Analysis

Patient and clinician data were analyzed separately using principles of codebook thematic analysis (Braun & Clarke, 2006). Consistent with QD and pragmatism, codebook thematic analysis organized patterns in participant accounts while maintaining a low-inference descriptive orientation. Interview transcripts were imported into and coded using ATLAS.ti version 24 software (ATLAS.ti Scientific Software Development GmbH, 2024). An expert qualitative researcher external to the research team and a study team member used analyst triangulation throughout the analysis to enhance rigor and credibility. The unit of coding was a meaningful segment of text conveying a discrete idea relevant to intervention refinement or assessment procedures. The analysts developed a codebook using deductive and inductive strategies: initial codes were derived from the interview guides and study aims, and additional codes were added during iterative analysis to capture unanticipated participant perspectives. The qualitative expert and study team member independently coded the transcripts and met weekly to compare coding, discuss discrepancies, refine code definitions, ensure consistent codebook application, and apply revisions to subsequently coded transcripts. Consensus discussions supported analytic rigor rather than serving as a quantitative measure of interrater reliability. Text segments were reviewed by code and summarized across transcripts; code summaries were then grouped into broader conceptual categories and synthesized into themes by two expert qualitative researchers and the study team member. Themes were identified based on recurrence across participants, salience within participant narratives, and relevance to the study’s intervention development questions (Braun & Clarke, 2006). Theme importance was not determined solely by frequency; themes were retained when they captured concepts highly relevant to intervention refinement, participant experience, or the future feasibility study, even when discussed by fewer participants.

Following theme development, findings were compared across groups to identify areas of convergence, divergence, and complementary perspectives regarding intervention format and content, recruitment strategies, and assessments.

### 2.10. Sample Adequacy

Interviews were analyzed concurrently with data collection. The coding team met regularly to review emerging codes and themes and to assess whether additional interviews were generating substantively new information relevant to the study aims. Because the study had a focused objective of refining components of the proposed DBT-ST intervention rather than developing a novel explanatory theory, sample adequacy was guided by principles of information power. Concurrent analysis suggested thematic stabilization, and the sample provided sufficient information power for these focused intervention-refinement questions. Later interviews largely reinforced existing concepts and did not substantially alter the developing thematic framework.

### 2.11. Transparency and Openness

Interview guides are available in the Supplementary Materials. De-identified analytic materials, including codebooks, coding frameworks, and thematic summaries, are available from the corresponding author upon reasonable request, subject to IRB requirements, participant-consent provisions, and applicable privacy regulations. The qualitative interview data generated during this study contains sensitive information regarding participants’ cancer experiences, smoking behaviors, psychological symptoms, and personal health-related experiences. Therefore, full interview transcripts are not publicly available because complete de-identification cannot be guaranteed and public sharing could compromise participant confidentiality

## 3. Results

### 3.1. Study Sample

Eight patient participants agreed, consented, and completed the interviews. Two patients were interested and were lost to contact, three additional patients consented then declined to be interviewed for personal reasons, and another patient consented and then was lost to contact. No patients were excluded. Due to the large number of providers able to offer enrollment in this study, the total number of patients that were potentially approached is not available. The study team emailed 13 clinicians from diverse professions to recruit them to the study, nine of whom completed the study and four who did not respond to recruitment efforts. The clinician sample included three medical oncologists, two radiation oncologists, an advanced practice clinician, an oncology nurse, and two psychosocial oncology therapists. See **Table 3** for complete demographic data.

Interviews were completed from May 2024 to February 2025. For patient participants, two chose to conduct the interview by video call and six by telephone call. Patient interviews lasted about 30 to 70 minutes, averaging 47 minutes. For clinician participants, seven of the clinicians participated via video call and two via telephone call. Clinician interviews lasted from 27 to 58 minutes, averaging 40 minutes.

### 3.2. Participant Perspectives on the Proposed Intervention Components

Across the patient and clinician datasets, participants viewed several features of the proposed intervention components as potentially useful, particularly opportunities for peer support and the teaching of concrete skills for responding to smoking triggers, distress, and difficult emotions. However, perspectives were conditional rather than uniformly favorable. Group participation could provide connection or accountability but could also be uncomfortable for people reluctant to disclose in front of others. Virtual delivery could reduce travel and scheduling burdens while creating technology and engagement challenges for some patients.

Similar tensions arose regarding the inclusion of patients not ready to quit, the fit of mindfulness with patients’ beliefs, and the burden of the proposed assessment battery. **Tables 5 and 6** present the complete theme summaries and additional quotations from patient and clinician participants respectively; selected exemplars are integrated below.

#### 3.2.1. The Group Format Offered Support but Would Not Suit Every Patient

Patients and clinicians both identified potential mechanisms through which a group could support smoking behavior change and offer other benefits. These included learning from others, receiving encouragement, recognizing shared experiences, and developing accountability. One patient explained, “As far as being in a group, you can support each other as far as people giving suggestions on what they’re doing versus others” (PA17). Clinicians similarly anticipated reciprocal learning, with one describing how patients further along in the process could model change for others: “The patients learn from one another, and they receive the support. The folks who are farther down the process are able to model for people who are less far down the process. There’s just a lot of great reciprocal learning there” (C05).

A contrasting patient perspective illustrated how discomfort with groups or interpersonal participation could limit engagement: “Because for the simple fact, I hate people. I hate crowds…I would be one of those people that sit there and don’t say anything” (PA13). This negative case directly articulated the personal unease that the group format could produce for some patients.

Participants also considered whether variation in cancer stage would strengthen or complicate group cohesion. Patients described interest in learning from people who had completed treatment, while clinicians anticipated that differences in treatment status could expose participants to useful perspectives and different smoking triggers. At the same time, one clinician raised a minority view that exposure to someone doing substantially better with cancer-related outcomes could generate resentment, jealousy, or unfavorable comparison rather than hope. Hearing about another patient’s success “could either work really, really well for self-motivation versus set up a part for pity versus jealousy…It could be a mixed package” (C09).

#### 3.2.2. Virtual Delivery Increased Access but Introduced Patient-Specific Barriers

Patients emphasized the convenience of virtual participation, particularly when balancing the intervention with medical appointments. PA04 stated, “I prefer that a lot of times ‘cause I’m on the go so much with other appointments that it’s just easier to fit it in that way.” Clinicians also recognized that virtual delivery could reduce travel, time away from work, and the burden of driving and parking. In this respect, patients and clinicians converged on virtual delivery as a potential way to make participation more manageable within the demands of cancer care.

Clinicians, but not patients, raised conditions under which virtual delivery might decrease rather than increase access. Their concerns focused particularly on older, rural, and lower-income patients and patients with less familiarity with video technology. One clinician noted that the older patients in their lung cancer practice “tend to strongly oppose anything virtual, anything like a video type. They like to do things in person” (C01). Clinicians also anticipated reduced attention and rapport online. C06 explained, “When you’re on a screen, it’s easier to get distracted…if I’m in the middle of doing something else, not really being as fully engaged.”

#### 3.2.3. Sessions Need to be Shorter

Both groups questioned whether 90-minute sessions would be manageable. Patients connected session duration to treatment demands and limited energy; PA17 suggested shortening sessions because patients’ “time is precious.” Clinicians focused on attention and retention, with C01 arguing that the material should be “cut down dramatically to 60 minutes” if patients were expected to retain and use it. Patients and clinicians also emphasized that attendance would depend on treatment schedules and whether participants felt well enough to attend. These shared concerns indicated the intervention needed to fit fluctuating health, energy, concentration, and treatment demands.

#### 3.2.4. Readiness to Quit Created a Tension Between Engagement and Reach

Views differed regarding the inclusion of patients who were not ready to quit smoking. A supportive clinician perspective framed participation as a pathway toward readiness rather than something that should follow an established commitment to quit: “You including them is not going to hurt them. All it can do is provide them with additional tools that may get them to the point at which they then are ready to quit” (C03). From this perspective, exposure to skills and peers committed to cessation could help motivation develop over time.

The patient data included a contrasting concern about whether people without current quit intentions would enroll or remain engaged: “I doubt they would [commit]. If they’re not ready to quit, why would they join a quitting [study]?” (PA14). Clinicians likewise identified low motivation and denial as possible barriers, although they did not view those barriers as reasons for exclusion. Thus, the data presented readiness to quit as an unresolved recruitment and participation consideration rather than consensus in favor of inclusion.

#### 3.2.5. Skills Were Perceived as Relevant, but Their Fit Depended on Presentation

Patients and clinicians commonly described DBT skills of mindfulness, emotion regulation, and distress tolerance as potentially relevant to smoking and cancer-related distress. Patients described mindful breathing, pausing, and distraction from distress as calming themselves or interrupting smoking-related reactions. For example, PA18 said, “Whenever I get upset, I pause. I take a deep breathe…and rationale the situation of what upset me and why I’m upset.” Clinicians similarly described DBT skills as a way to more easily recognize triggers and create space between an urge and the decision to smoke. These accounts suggest that the perceived credibility of DBT-ST skills rested on their practical use in interrupting established smoking patterns and managing cravings.

Regarding mindfulness specifically, a few clinicians raised concerns that the language of mindfulness could be unfamiliar, overly abstract, or inconsistent with some patients’ faith-related understandings. C05 explained that patients may “bristle with the term ‘mindfulness’” and worry “that that’s more of a religious thing rather than a mental health thing.” Another clinician suggested that acceptance might depend on hearing that mindfulness had worked for peers.

Accordingly, the findings identified an anticipated clinician concern that acceptability may depend on concrete explanation, language choice, and social validation.

Participants differed in how strongly they associate emotions with smoking. PA19 described identifying the emotional pathway to smoking as useful: “If I can get the emotion part of it down, because if one thing is stressed out, start smokin’ and then, ‘Oh, okay, well, what drove me to this point?’ Bein’ able to identify that within myself…I think more actionable steps to prevent goin’ there.” In contrast, PA10 stated, “I don’t let emotion get involved with me at all,” describing a factual rather than emotional approach. This negative case suggests that emotion regulation content may resonate most directly with patients who recognize an emotion–smoking connection, whereas others may require a more behaviorally or practically framed rationale.

#### 3.2.6. Patients and Clinicians Viewed Assessments from Different Vantage Points

Patients found the proposed assessments understandable and relevant to their experiences of cancer, depression, anxiety, and coping. PA18 commented, “I thought those questions were really good questions…kept asking me that question in a different way.” Clinicians also regarded the assessed domains as relevant to the proposed intervention. Thus, the groups converged on the conceptual relevance of the measures.

Their perspectives diverged regarding anticipated burden. Clinicians questioned whether medically burdened patients could sustain attention across the complete battery and warned that fatigue might reduce data quality. C06 stated, “It’s a lot. It’s long, so that’s a little bit concerning…people do get tired and just start rushing through them versus really thinking about that too is another question.” Patient feedback uniformly reflected confidence they could complete the measures, whereas clinician feedback reflected concerns about how the battery might function across patients experiencing illness, treatment effects, or fatigue.

#### 3.2.7. Engagement Depended on Fit with Cancer Care

Patients linked engagement in the intervention to scheduling and feeling well enough, and clinicians similarly anticipated missed sessions when patients were ill, receiving treatment, or moving between facilities. Clinicians also recommended that recruitment extend beyond physician referral, although both patients and clinicians supported endorsement from a trusted healthcare professional as lending the intervention credibility.

### 3.3. Conclusion

Collectively, these findings indicated that anticipated success of the intervention was conditional on flexible delivery, manageable session length, accessible materials, and accommodation of fluctuations in health and treatment burden, beyond simple interest in the program.

## 4. Discussion

This qualitative descriptive study of components of our proposed DBT-ST intervention for cigarette smoking by patients with cancer resulted in valuable feedback from participants. They strongly supported some components of the intervention, such as teaching patients DBT-ST skills for being more present, managing emotions, and dealing with stress, while sharing mixed opinions on other components, such as its virtual format and inclusion of patients not ready to quit. Perceived benefits extended beyond smoking cessation to improved quality of life, enhanced social support, decreased anxiety and depression, and acquisition of skills universally applicable to managing life’s challenges, including cancer.

A cross-cutting theme is that patients felt more capable and less burdened regarding virtual technology and assessments than clinicians perceived they would. Patients were uniformly supportive of virtual sessions for accessibility and ease of scheduling. Clinicians were concerned that their older, rural, and lower-income patients would struggle with video technology or that patients would become disengaged during sessions; however, no patients spoke of technology or disengagement during sessions as concerns. Regarding assessments, patients had positive feedback and expressed no concerns, while clinicians thought patients might find them confusing, repetitive, or too much of a time burden. Assessments would be used in future trials, but not clinical practice, and given that patients did not feel these to be a burden, the decision was made to consider them as outcome measures in the future feasibility study.

Our results aligned with recent research into cessation interventions for patients with cancer. Shenton, et al. interviewed patients, clinicians, and cessation program staff to discover barriers and facilitators to smoking cessation in a single cancer center (Shenton et al., 2023). They found that patients endorsed interventions focused on stress management to enable quitting, and they recommended that cessation services tailored to patients with cancer be offered throughout treatment and survivorship. These results suggest potential support for DBT-ST distress tolerance skills. Charlot, et al. conducted a mindfulness-based group cessation intervention for patients with cancer from low socioeconomic and minority populations (Charlot et al., 2019). The authors reported that participants reduced cigarette use by 41% on average, were satisfied with the program, and made positive lifestyle changes. While this population is a subset of our population, the results suggest that the mindfulness component of DBT-ST may help for patients with cancer. Mindfulness is known to improve tobacco cessation in the general population and needs further assessment in patients with cancer who smoke (Oikonomou et al., 2017; Spears et al., 2017; Vidrine et al., 2016).

### 4.1. Intervention Implications

Given the results of this qualitative study, several modifications to the intervention were made. **Table 6** links participant feedback to the resulting changes to our DBT-ST intervention.

### 4.2. Limitations

Our study’s small sample size and composition limited the diversity of perspectives and generalizability. Patient participants were mostly male and did not include persons of Hispanic or Latino ethnicity or Asian racial identity. Clinician participants were mostly female and White. Thus, important sex, racial, and ethnic perspectives are missing from the analyses, which inhibits the transferability of findings to other cancer and clinician populations. Other limitations include recruitment from only one cancer center in the southeastern United States which suggests a regional bias, and the potential for bias in favor of the intervention due to three members of the intervention design team also conducting interviews. Another limitation is that we included patients who smoked and had quit but did not separately analyze responses from the two groups due to sample size limitations. Future studies with larger samples could include sub-analyses by patient smoking status to assess differences in perspectives and concerns.

Several interviewers were members of the intervention development team who held favorable expectations regarding DBT-ST. Participants were aware that interviewers were affiliated with the project and patient recruitment occurred through clinical teams, creating potential for social desirability bias and increased endorsement of intervention components. Although efforts were made to elicit both positive and negative perspectives and to incorporate external qualitative review, findings should be interpreted with recognition that interviewer affiliation may have influenced participant responses.

### 4.3. Transferability

Transferability should be considered in light of the characteristics of the study sample. Although participants represented diverse racial and sex backgrounds, experiences may differ according to cancer type, disease stage, treatment status, and symptom burden. Similarly, perceptions of the intervention may vary by smoking status, nicotine dependence, quitting history, readiness to quit, and time since smoking cessation. The present study was not designed to examine differences across these characteristics. Future research with larger and more diverse samples should explore how intervention experiences and outcomes may vary across cancer- and tobacco-related subgroups.

### 4.4. Future Feasibility Study

This study focused on intervention development and stakeholder feedback rather than intervention delivery. Consequently, several research design and participant safety considerations remain for planned feasibility testing. Future work will establish and evaluate procedures related to facilitator training and supervision, treatment fidelity, participant safety monitoring, adaptation of specific DBT skills for medically vulnerable oncology populations, management of acute psychological distress, group confidentiality protections, and integration with pharmacotherapy and other aspects of comprehensive oncology tobacco treatment.

Although clinicians expressed concerns regarding burden despite uniform support from patients for the assessments, the retained measures capture key constructs targeted by the intervention, including tobacco use, emotion regulation, distress tolerance, psychological symptoms, and cancer adjustment. Because this study focused on intervention development rather than finalization of a feasibility trial protocol, we elected to retain all candidate measures pending future evaluation of participant burden, completion rates, and measure performance.

## 5. Conclusion

These findings suggest that key components of our DBT-ST intervention may support smoking cessation among patients with cancer and warrant evaluation in a feasibility trial. We will continue to develop our DBT-ST intervention for the oncological patient population using the NIH Stage Model (Onken et al., 2014) to determine its feasibility, efficacy, effectiveness, and sustainability.

## Data Availability

All data produced in the present study are available upon reasonable request to the authors

## Acknowledgments

The authors would like to acknowledge and thank James Morgan, Thomas Lycan, Jr., DO, Michael Farris, MD, Ryan Hughes, MD, William J. Petty, MD, and the Psychosocial Oncology Program therapists for recruiting patients to this study.

## Statements and Declarations

### Ethical Considerations

This study was approved by the Institutional Review Board of Advocate Health – Wake Forest University School of Medicine on March 23, 2024.

### Consent to Participate

All participants provided verbal informed consent prior to enrollment in the study.

### Consent for Publication

not applicable

### Declaration of Conflicting Interest

We have no known conflicts of interest to disclose.

### Funding Statement

Research supported in part by the Qualitative and Patient-Reported Outcomes Shared Resource, the Tobacco Control Center of Excellence, and the Cancer Prevention and Control Program of the Atrium Health Wake Forest Baptist Comprehensive Cancer Center’s NCI Cancer Center Support Grant P30CA012197.

### Data Availability

Comprehensive reports of patient and clinician thematic analyses are available from the corresponding author upon request.

## Notes

### Competing Interest Statement

The authors have declared no competing interest.

### Author Declarations

The Institutional Review Board of the Wake Forest University School of Medicine gave ethical approval for this work.

### Summary of Updates

Abstract, main body of manuscript; and tables have been revised

