## Supplementary material for "A Dialectical Behavior Therapy – Skills Training Intervention for Cigarette Smoking by Patients with Cancer: Qualitative Descriptive Study of Intervention Components Prior to Feasibility Trial": Table 1

**Table 1.** *Adaptation of Dialectical Behavior Therapy – Skills Training (DBT-ST) for Cigarette Smoking by Patients with Cancer*

| **Intervention Structure**   - Eight 90-minute sessions, one per week across eight weeks - Group intervention conducted virtually - Led by facilitator with DBT-ST training from Behavioral Tech Institute* - 6-10 patients with cancer per group - Patients could be from any stage of the cancer journey, with interest in quitting smoking ranging from ready to not ready - Use of nicotine replacement or other smoking cessation aids recommended at first session but not required** | | |
| --- | --- | --- |
| **Intervention Content by Sessions** | **Key Reasons Why Patients with Cancer Smoke** | **DBT-ST Skills to Address Smoking and Cancer** |
| 1 – Orientation | Fatalistic beliefs, hopelessness about the future  Environmental and event triggers | *Goal setting*: create a meaningful “life worth living” with cancer  *Chain analysis*: identify vulnerabilities to smoking, triggering events, thoughts and feelings leading to smoking, consequences |
| 2 and 3 – Mindfulness | Smoking as an automatic, entrenched, lifelong habit for coping with adversity such as cancer | *Mindful practices*: become aware of fluctuating emotions and runaway thoughts that come with cancer without self-judgment. Realistically assess difficulties and learn not to overreact; tolerate urges to smoke without acting  *Wise Mind*: integrate logic and emotions to decide how to handle life’s disturbances without smoking; trust inherent wisdom in self |
| 4 and 5 –  Distress Tolerance | Overwhelming situations and unbearable cravings | *Crisis survival skills:* strategies for coping with severe discomfort and learning to self-soothe instead of smoking  *Radical acceptance*: while not liking one’s reality, fully accepting distressing truths of life with cancer, constant change, hard decisions, and difficulties with quitting smoking |
| 6 and 7 –  Emotion Regulation | Intense negative emotional states – depression, anxiety, stress – from dealing with cancer, other life events, or withdrawing from smoking | *Understand and name emotions*: learn the role of emotions and which emotions are present and threaten to overwhelm  *Change emotions*: instead of smoking, “check the facts” of evidence for thoughts and feelings, take “opposite action” to emotional urges, “problem solve” solutions to situations, engage in “pleasant activities” |
| 8 - Closing | Unrealistic plans for quitting and not relapsing | *Dialectical abstinence*: “Wise Mind” abstinence plan accounting for overwhelming emotions and unexpected events, plan for reducing harm of relapse if it occurs |

**Behavior Tech Institute was founded by the creator of DBT, Marsha Linehan, and is a widely recognized organization for DBT training and implementation programs*

***Use of cessation aids will be tracked in future clinical trials*
