## Supplementary material for "A Dialectical Behavior Therapy – Skills Training Intervention for Cigarette Smoking by Patients with Cancer: Qualitative Descriptive Study of Intervention Components Prior to Feasibility Trial": Table 2

**Table 2***. Assessments Presented for Participant Feedback*

| **Assessment** | **Items** | **Estimated Completion Time** |
| --- | --- | --- |
| Cancer Patient Tobacco Use Questionnaire (C-TUQ)^40^ | 22 | 5 min |
| Difficulties in Emotion Regulation Scale (DERS)^41^ | 36 | 5-10 min |
| Distress Tolerance Scale (DTS)^42^ | 15 | 1-3 min |
| Mental Adjustment to Cancer (MAC)^43^ | 40 | 10-15 min |
| Depression, Anxiety and Stress Scale (DASS-21)^44^ | 21 | 3-10 min |
