## Supplementary material for "A Dialectical Behavior Therapy – Skills Training Intervention for Cigarette Smoking by Patients with Cancer: Qualitative Descriptive Study of Intervention Components Prior to Feasibility Trial": Table 3

**Table 3.** *Sociodemographic Characteristics of Participants*

| Characteristics of Participants | Patient | | Clinician | | Total Sample | |
| --- | --- | --- | --- | --- | --- | --- |
|  | *n* | *%* | *n* | *%* | *n* | *%* |
| Sex  Male  Female  Age in Years  Average Age  Age Range  Race  Asian  Black  White | 6  2  60.6  51-80  2  6 | 75  25  25  75 | 2  7  44.9  37-62  1  8 | 22  78  11  89 | 8  9  52.3  37-80  1  2  14 | 47  53  6  12  82 |
| Interview Format |  |  |  |  |  |  |
| Video interview | 2 | 25 | 7 | 78 |  |  |
| Audio interview | 6 | 75 | 2 | 22 |  |  |
| Patient Participant Cancer Type |  |  |  |  |  |  |
| Small cell lung carcinoma | 3 | 37.5 |  |  |  |  |
| Non-small cell lung carcinoma | 1 | 12.5 |  |  |  |  |
| Lung adenocarcinoma | 2 | 25 |  |  |  |  |
| Myelodysplastic syndrome | 1 | 12.5 |  |  |  |  |
| Thyroid carcinoma | 1 | 12.5 |  |  |  |  |
| Patient Participant Smoking Status |  |  |  |  |  |  |
| Currently smoking | 3 | 37.5 |  |  |  |  |
| Previously smoked | 5 | 62.5 |  |  |  |  |
| Average cigarettes per day* | 22.1 |  |  |  |  |  |
| Median cigarettes per day* | 15 |  |  |  |  |  |
| Range of cigarettes per day* | 5 - 60 |  |  |  |  |  |
| Average quit time in weeks** | 22.3 |  |  |  |  |  |
| Median weeks quit** | 6 |  |  |  |  |  |
| Range in weeks of quit time** | 2 - 86 |  |  |  |  |  |
| Participants using nicotine replacement therapy** | 2 | 25 |  |  |  |  |
| Clinician Participant Professions  Medical oncologist  Radiation oncologist  Oncology physician assistant  Oncology nurse navigator  Psychosocial oncology therapist |  |  | 3  2  1  1  2 |  |  |  |

*Includes participants who were currently smoking and those who had quit smoking. Data missing for one participant.

**Includes only participants who quit smoking.
