## Supplementary material for "A Dialectical Behavior Therapy – Skills Training Intervention for Cigarette Smoking by Patients with Cancer: Qualitative Descriptive Study of Intervention Components Prior to Feasibility Trial": Table 4

**Table 4.** *Summary of Patient Qualitative Results by Intervention Component*

| **Component** | **Summary of Patient Perspectives** | **Illustrative Quotation** |
| --- | --- | --- |
| **Group format and peer support** | Patients described opportunities for peer support, shared experiences, practical suggestions, and accountability. Group participation was not universally appealing; discomfort with groups or speaking in front of others could limit engagement. | “As far as being in a group, you can support each other as far as people giving suggestions on what they’re doing versus others.” (PA17) |
| **Group composition** | Patients anticipated potential value in interacting with people at different stages of cancer treatment or recovery and learning from their experiences. | “…those who beat cancer and recovering, to know what their thoughts are and how they feel after treatment and how they dealing and how life is for them, would be interesting to me.” (PA18) |
| **Virtual delivery** | Virtual participation was viewed as convenient and potentially easier to accommodate alongside cancer treatment and other medical appointments. | “I prefer that a lot of times ‘cause I’m on the go so much with other appointments that it’s just easier to fit it in that way.” (PA04) |
| **Readiness to quit smoking** | Patients lacked consensus on inclusion of people not ready to quit smoking. | “I doubt they would [commit]. If they’re not ready to quit, why would they join a quitting [study]?” (PA14) |
| **Mindfulness and distress tolerance skills** | Patients described mindfulness, breathing, pausing, distraction, and related strategies as potentially useful for managing distress and smoking triggers. | “Now, I think that would be something good to go over because the pros and cons of the distress, especially the distress, because most of the time when you get stress out, that’s when you’re – you do your most craving on anything.” (PA15) |
| **Emotion regulation skills** | Some patients recognized connections among emotions, stress, and smoking and viewed emotion-regulation strategies as potentially useful. Others did not identify emotions as central to their smoking or coping. | “I don’t let emotion get involved with me at all…I knew I had problems, I knew I had cancer, but that’s the biggest emotion of all and it shut every else down and just had to go with the factual end of it…I didn’t really worry about the emotional part of it.” (PA10) |
| **Assessments** | Patients generally described the proposed assessment content as understandable and relevant to their experiences with cancer, emotional health, and coping. | “I thought those questions were really good questions…kept asking me that question in a different way.” (PA18) |
| **Recruitment and engagement** | Participation was viewed as dependent on fitting sessions around cancer treatment, medical appointments, fatigue, and periods of feeling unwell. Longer sessions could be difficult for some patients. | “I think the rate of completion may be lower just due to timeframe and committing to the meetings and the process. I think these patients are …busy with life, work, and cancer treatment. We’re adding two, three hours a week to their already busy schedule, so I think participation would be limited more to the duration of the treatment, but I think there would be a lot of interest in this.” (PA11) |
| **Overall conclusion** | Skills can help regulate emotions, be used as replacements or distractions from smoking. | “Especially with the anxiety. The distracting yourself and the breathing. The depression, of course, if you’re dwelling on something and you start doing something else to distract yourself, a lot of times it’ll lessen.” (PA04) |

**Note.** PA = patient participant. Quotations are reproduced verbatim except where ellipses indicate omitted text and brackets indicate clarification.
