## Supplementary material for "A Dialectical Behavior Therapy – Skills Training Intervention for Cigarette Smoking by Patients with Cancer: Qualitative Descriptive Study of Intervention Components Prior to Feasibility Trial": Table 5

**Table 5.** *Summary of Clinician Qualitative Results by Intervention Component*

| **Component** | **Summary of Clinician Perspectives** | **Illustrative Quotation** |
| --- | --- | --- |
| **Group format and peer support** | Clinicians anticipated that groups could provide support, normalization, reciprocal learning, and opportunities for participants to model behavior change for one another. | “I think when you have group interventions and people talking about things and shared experiences and being brutally honest about what the rate-limiting barriers to smoking cessation are, it goes around the room. People can have a kindred spirit and be supportive of one another and cheer each other on.” (C09) |
| **Group composition** | Variation in cancer stage, treatment status, and cessation progress could expose participants to useful perspectives. Clinicians also noted that differences among participants could sometimes produce unfavorable comparisons rather than motivation. | “If I’m not doing well, would I want to see somebody in the group that’s stage 1 and hunky-dory and living life to the fullest? I personally would maybe have a little resentment toward them, but I don’t know.” (C08) |
| **Virtual delivery** | Clinicians identified potential advantages of virtual delivery, including reduced transportation, parking, scheduling, and work-related burdens. They also anticipated technology barriers, preference for in-person interaction, and reduced engagement for some patients. | “One plus is I think it’ll allow more patients to be able to be part of it, being more accessible, especially if they would have to travel to the hospital or anything.” (C07) |
| **Session duration and engagement** | Clinicians questioned whether 90-minute sessions would be manageable given treatment schedules, illness, fatigue, attention, and competing demands. Shorter sessions were suggested as potentially more manageable. | “If you’re teaching for 90 minutes, people are not gonna retain that because it’s just too much stuff, so whatever you’re teaching them – if you want them to retain stuff and actually use it, whatever you’re teaching them needs to be cut down dramatically to 60 minutes.” (C01) |
| **Readiness to quit smoking** | Clinicians suggested that patients who were not yet ready to quit might still benefit from exposure to cessation skills and peer experiences that could increase subsequent motivation. Low motivation could nevertheless affect engagement. | “You including them is not going to hurt them. All it can do is provide them with additional tools that may get them to the point at which they then are ready to quit.” (C03) |
| **Mindfulness skills** | Mindfulness was viewed as potentially useful for recognizing triggers and interrupting automatic responses to smoking urges. Clinicians emphasized the importance of accessible terminology and anticipated that the concept could be unfamiliar or raise faith-related concerns for some patients. | “I feel like most people need more mindfulness in their life from day one to death. I think that from my experience with patients, the majority of the decisions they make around smoking are not rational…Even the impulse to quit cold turkey, even if they do it’s not rational in any sense. I think that the ability to take a step back and conceive of your own situation in a more objective format with some level of impartiality would be very helpful.” (C03) |
| **Emotion regulation and distress tolerance skills** | Clinicians viewed emotion-regulation and distress-tolerance skills as relevant to cancer-related distress and smoking triggers and recommended presenting skills concretely and accessibly. | “A lot of times, patients go for that cigarette because they don’t have the ability to self-soothe, or they don’t have the ability to manage or related to that strong emotion without automatically going and lighting up. Getting them to pause for a second, recognize the urge, do something different, and then have the coping skills to be able to ride that wave of that urge until it passes is gonna be key to facilitate that new learning and show them that they don’t always have to jump to the cigarette in order to cope.” (C05) |
| **Assessments** | Clinicians considered the assessment domains relevant but expressed concern about the length of the battery, fatigue, attention, and the possibility that participants might rush responses. | “It’s a lot. It’s long, so that’s a little bit concerning…people do get tired and just start rushing through them versus really thinking about that too is another question.” (C06) |
| **Recruitment** | Clinicians suggested that endorsement from trusted healthcare professionals could increase credibility but recommended multiple recruitment pathways rather than relying exclusively on physician referral. | “Definitely providers-as patients come into clinic-providers mentioning it, some resources in our clinic rooms or having some of our staff help pass out information about it and then the provider just mentioning, ‘Hey, this is a great program, I recommend it for you.” (C11) |

**Note.** C = clinician participant. Quotations are reproduced verbatim except where ellipses indicate omitted text and brackets indicate clarification.
