## Supplementary material for "A Dialectical Behavior Therapy – Skills Training Intervention for Cigarette Smoking by Patients with Cancer: Qualitative Descriptive Study of Intervention Components Prior to Feasibility Trial": Table 6

**Table 6.** *Intervention Modifications from Participant Results*

| **Topic Area** | **Original Intervention** | **Participant Results** | **Revised Intervention** |
| --- | --- | --- | --- |
| Intervention Structure | 90-minute sessions | Sessions are too long | Reduce to 60-minute sessions |
| Intervention Structure | Virtual group format | Mixed supportive and critical themes, but more supportive. Concerns about technological barriers | Retain virtual group format but will have a staff member at each session dedicated to helping participants successfully join the group online |
| Intervention Structure | Inclusion of patients from any stage of the cancer journey, with interest in quitting smoking ranging from ready to not ready | Mixed supportive and critical themes, but more supportive | Retain inclusion criteria at present; re-evaluate after feasibility study |
| Intervention Structure | N/A | Clinicians spontaneously suggested ideas for intervention structure | Ensure skills training balances didactic teaching and opportunities for group discussion and engagement |
| Intervention Content | Teaching dialectical behavior therapy (DBT) skills as originally designed | Adapt skills and teaching style to be more concrete and accessible to all literacy levels, cultures, and belief systems, especially with mindfulness skills | Revise language, framing of concepts, and instructions to make the intervention concrete, value-neutral, and easy to understand |
| Intervention Content | N/A | Patients spontaneously suggested ideas for intervention content | Ensure skills training incorporates smoking cessation barriers and facilitators identified by patients, e.g., managing to be around people who smoke, seeing smoke, taking advantage of cancer as a wake-up call, dealing with guilt |
| Assessments | Six patient-reported assessments evaluating smoking, psychological symptoms, and dialectical behavior therapy – skills training (DBT-ST) skills | Mixed themes, with patients finding assessments easy to understand and relevant, but clinicians concerned about excess length and number of questions | Based on patient supportive feedback, and relevance to clinical and research outcome assessment, retain assessments for ongoing consideration as outcome measures in feasibility trial |
| Recruitment and Participation | N/A | With cancer population, challenges with busy schedules and of participants feeling well enough to attend | Offer sessions in late afternoons or evenings to accommodate complex patient schedules  Offer session materials to participants who must miss due to emergencies or illness |
| Recruitment and Participation | N/A | Clinicians recommending study to patients is most effective method, but use other recruitment pathways too | Recruit using both clinician-to-patient and direct-to-patient (e.g., patient portal) methods |
