## Supplementary material for "A Dialectical Behavior Therapy – Skills Training Intervention for Cigarette Smoking by Patients with Cancer: Qualitative Descriptive Study of Intervention Components Prior to Feasibility Trial": S3

**S3. Patient Interview Guide**

**Introduction**

Welcome and thank you for meeting with me today. Our meeting today will cover two topics. First, we will discuss your thoughts on the questionnaires you took online. Second, I will describe the program we developed to help patients with cancer reduce or quit smoking and ask for your thoughts. The basis for this program, Dialectical Behavior Therapy or DBT, was described in the Information Sheet you read prior to verbally agreeing to participate in this study. Then I would like to discuss your thoughts on it. Our interview should take about one hour. Before we begin, do you have any questions?

**Patient Questionnaires**

We are looking at several factors to better understand how to help patients with cancer reduce or quit smoking through our program. These factors include depressive and anxious feelings, stress levels, the ability to tolerate and manage distressing emotions and stress, and the mental adjustment to cancer. We also document tobacco use history to compare patient smoking behaviors to these factors. We are gathering this data using questionnaires, which we plan to use in future studies about our program to measure its effectiveness. The questionnaires we are using are the ones you took online before today’s meeting. For this portion of our discussion, I would like to find out more about your experience with these questionnaires. Before we move on, do you have any questions?

Questions

1. Tell me about your experience taking the questionnaires, especially being able to understand and answer the questions.
   a. What other thoughts or comments do you have about the experience of taking the questionnaires?
2. How are depressive and anxious feelings relevant to your cancer experience and to smoking?
3. How is stress relevant to your cancer experience and to smoking?
4. How is the ability to tolerate and manage distressing emotions and stress relevant to your cancer experience and to smoking?
5. How is the mental adjustment to cancer relevant to your cancer experience and to smoking?
6. What are some additional thoughts or feelings you have related to your experiences with cancer and continued smoking?

**Program**

Our virtual eight-session group program is designed for 6-10 patient participants, facilitated by one behavioral health professional such as an advanced practice provider, a licensed mental health provider, or a health educator. Patient participants can be at any stage of cancer and in any phase of treatment including completion. Patients will not be required to quit smoking to participate. Our eight group sessions will last ninety minutes each and consist of an introduction and orientation session, two sessions on mindfulness skills, two sessions on emotion regulation skills, two sessions on distress tolerance skills, and one session for reflection and planning for the future. Before I begin discussing these skills, do you have any questions?

Mindfulness skills

- These skills allow us to “press the pause button” when experiencing an upsetting event, thought, or feeling so that we are not overcome by it.
- By pausing and gaining understanding of our experience, we regain our power to choose healthy behaviors rather than act without thinking or become overwhelmed by what’s going on around and within us.
- Here are two examples of the mindfulness skills we plan on teaching:

*Wise Mind:* The Wise Mind skill is a way to stay balanced so that our decisions and behaviors reflect what we really want for ourselves. The Wise Mind has two parts – reasonable mind and emotional mind. Sometimes we think and act from our reasonable minds. We are rational, cool, and focused on what we need to do. An example is making a plan to quit smoking – we plan a quit date, predict what might trip us up, and decide how we will handle things in a rational, logical way. But sometimes we think and act from our emotional minds. We have strong feelings, get heated, and act according to our mood. An example is dealing with cigarette craving – we are overwhelmed with need and the fear that we can’t withstand the craving, then we smoke. When we think and act from Wise Mind, we value both reason and emotion, seeing the bigger picture. An example would be making a reasonable plan to quit smoking that also is very serious about preparing for the times when we are overwhelmed with emotions and in danger of relapsing. The Wise Mind skill is the most important skill in all of Dialectical Behavior Therapy.

*Expanding Awareness Exercise*: To help people stay in or get back to Wise Mind, we will teach daily practices that use their breathing to calm down and be more aware of what they are experiencing. Breathing practices need repetition to make them automatic for when people need to use them, so we will suggest doing them every day. People who smoke cigarettes are used to taking deep breaths, so sometimes breathing practices are easier for them. One example of a breathing practice we will teach is called the Expanding Awareness Exercise. In this exercise, when we breathe in, we focus our minds on our inner experiences, and when we breathe out, we keep that inward focus but also focus on our surroundings. We do this about ten times in a set and do a set one or more times per day. Through repetition, we develop increased awareness of our inner and outer experiences, which will help us make better decisions when challenges come.

Emotion regulation skills

- These skills help us manage our emotions and soothe the discomfort in our bodies during an emotionally intense moment without behaving hastily or inappropriately.
- They also help us refocus attention on a goal and continue moving toward it regardless of mood.
- Here are two examples of the emotion regulation skills we plan on teaching:

*Understanding and Naming Emotions*: We will help people gain greater emotional control through understanding and naming their emotions, which we hope will lead to less reliance on smoking for dealing with emotions. Dealing with cancer at any stage can involve a range of challenging and overwhelming emotions. To understand emotions, we need to know what they do, which is to motivate and prepare us for action, help us communicate with others, and give us important information about situations and about ourselves. Some reasons it’s hard to regulate emotions include not knowing how to do it and becoming too overwhelmed to use the skills we have. We can teach ourselves how to stop and name our emotions when we are feeling them, and to learn the many different words for the basic emotions of anger, disgust, envy, fear, happiness, jealousy, love, sadness, shame, and guilt. Through understanding and naming, we feel more in charge of our emotional experiences and are less likely to let emotions determine our actions.

*Accumulating Positive Emotions*: when people focus on accumulating positive emotions, they can build a life worth living in the long term. Doing so can help people with cancer have hope in the face of their disease and its treatment and perhaps rely less on smoking to feel better. We can take seven steps to accumulate positive emotions for the long term: get started, identify some of our most important values, choose one value to focus on, name some goals for living according to that value, choose one goal, list small actions we can take towards that goal, and take that action. An example would be to value relationships, have a goal to spend more time with a loved one, and taking action to schedule that time.

Distress tolerance Skills

- These skills help us make a challenging or painful situation more tolerable so that it is possible to refrain from impulsive actions that can make the situation worse.
- Here are two examples of the distress tolerance skills we plan on teaching:

*Pros and Cons*: We can make a pros and cons list to plan for times when we might become overwhelmed and take actions that make things worse, such as having an incredibly strong urge to smoke and giving in to that urge. We list our personal pros and cons of acting on that urge. A pro might be to feel great relief in the short term and a con might be to disappoint ourselves. We can also list our pros and cons of not acting on that urge. A pro might be satisfaction that we made a good choice for our future health and a con might be the distress from having to fight through the urge. Through planning via a pros and cons list, we can improve our chances of taking actions that improve our well-being when the urges hit.

*TIPP Skills*: When we feel great distress, we can use the TIP skills to change our body chemistry and reduce our distress. “T” stands for Tip the Temperature, meaning applying cold to our face such as an ice pack or putting our face in ice water. “I” means Intense Exercise – getting our heart rates up through running, fast walking, jumping, lifting weights, playing sports, etc. The first “P” stands for both Paced Breathing and the second “P” stands for Paired Muscle Relaxation. Paced Breathing is a technique to slow down our breathing and calm our nerves by deliberately inhaling slowly and exhaling even more slowly. Paired Muscle Relaxation involves tensing our bodies when we inhale and releasing the tension as we exhale. After any of these techniques, we take time to notice the change in our bodies and our distress levels.

Now that I have presented the program, I would like to discuss your thoughts on it.

Questions

First, let’s discuss the format.

Overview of format

1. What are your thoughts on:
   a. Using a virtual group program for patients with cancer?
   b. Including patients at any cancer stage or phase of treatment?
   c. Including patients not committed to quitting smoking?
   d. Conducting the group virtually?
   e. The number and length of sessions?
   f. Do you have any additional thoughts or suggestions on the group format?
2. To what degree would you find this group format helpful?
   a. Please rate the helpfulness of our suggested intervention format from 1-5 with 1 being unhelpful and 5 being very helpful.
3. To what degree would you find this group format practical and convenient?
   a. Please rate the practicality and convenience of our suggested intervention format for patients like you from 1-5 with 1 being not practical or convenient at all and 5 being highly practical and convenient.

Now let’s discuss the skills themselves.

DBT Skills

1. What do you think about the mindfulness skills we chose to include in our program?
2. What do you think about the emotion regulation skills we chose to include in our program?
3. What do you think about the distress tolerance skills we chose to include in our program?
5. How do you think these skills would help you mentally adjust to cancer?
6. What other thoughts or comments do you have on these skills?
7. Please rate the helpfulness of our suggested DBT skills content for you from 1-5 with 1 being unhelpful and 5 being very helpful.
8. Please rate the practicality of our suggested DBT skills content for you from 1-5 with 1 being impractical and 5 being highly practical.

**Recruitment, Participation** & **Retention**

The last topic related to the intervention that I would like to discuss with you is the likelihood that you would want to participate in this program and complete it.

1. What are your thoughts on participating in and completing this type of program?
2. What, if anything, discourages you from participating and/or staying in the program?
3. How can we encourage participation and completion of this program?
4. What suggestions for advertising would you recommend for reaching other patients who may be interested in reducing or quitting smoking?
5. What other thoughts or suggestions do you have for us?

**Closing**

Thank you very much for meeting with me today to discuss this important topic and for contributing to our efforts to develop an effective program for patients with cancer to cut down on or quit smoking.
