## Supplementary material for "A Dialectical Behavior Therapy – Skills Training Intervention for Cigarette Smoking by Patients with Cancer: Qualitative Descriptive Study of Intervention Components Prior to Feasibility Trial": S4

**S4. Clinician Interview Guide**

**Introduction**

Welcome and thank you for taking time out of your busy schedule to meet with me. Our meeting today will cover two topics. First, I will describe the behavioral intervention we developed to help patients with cancer reduce or quit smoking and ask for your thoughts. The basis for this intervention, Dialectical Behavior Therapy or DBT, was described in the Information Sheet you read prior to verbally agreeing to participate in this study. Second, I will ask for your thoughts on the patient questionnaires you reviewed in REDCap prior to our interview. Our interview will last about one hour. Before we begin, do you have any questions?

**Intervention**

Our intervention consists of eight ninety-minute virtual sessions delivered in a group setting. During the sessions, group members learn DBT skills from a behavioral health professional to help them reduce or quit smoking. Group members also gain support from one another. The DBT skills include mindfulness, emotion regulation, and distress tolerance. I will briefly present an overview of the intervention format, describe the DBT skills, and provide examples of how the skills will be taught to patients. Then I will ask for your opinions and perspectives.

Overview of format

Our group intervention is designed for 6-10 patient participants, facilitated by one behavioral health professional such as an advanced practice provider, a licensed mental health provider, or a health educator. The patients can be at any stage of cancer and in any phase of treatment including completion. Patients will not be required to quit smoking to participate. Our eight 90-minute virtual group sessions consist of an introduction and orientation session, two sessions on mindfulness skills, two sessions on emotion regulation skills, two sessions on distress tolerance skills, and one session for reflection and planning for the future.

**Questions**

Overview of format

1. What are your thoughts on:
   a. Using a group intervention for patients with cancer?
   b. Including patients at any cancer stage or phase of treatment?
   c. Including patients not committed to quitting smoking?
   d. Conducting the group virtually?
   e. The number (eight) and length (ninety minutes) of sessions?
   f. Do you have any additional thoughts or suggestions on the group format?
2. To what degree do you think patients like yours would find this group format helpful?
   a. Please rate the helpfulness of our suggested intervention format for patients from 1-5 with 1 being unhelpful and 5 being very helpful.
3. To what degree do you think patients like yours would find this group format practical and convenient?
   a. Please rate the practicality and convenience of our suggested intervention format for patients from 1-5 with 1 being not practical or convenient and 5 being highly practical and convenient.
4. How might patients from differing races/ethnicities, geographic locations, socioeconomic status, and/or educational levels find this group format helpful or not?
5. How might patients from differing races/ethnicities, geographic locations, socioeconomic status, and/or educational levels find this group format practical and/or convenient, or not?

DBT Skills

1. How might mindfulness skills help your patients reduce or quit smoking?
2. How might emotion regulation skills help your patients reduce or quit smoking?
3. How might distress tolerance skills help your patients reduce or quit smoking?
4. Do you have any additional thoughts or suggestions on the DBT skills we described?
5. Please rate the helpfulness of our suggested DBT skills content for patients with cancer from 1-5 with 1 being unhelpful and 5 being very helpful.
6. Please rate the practicality of our suggested DBT skills content for patients with cancer from 1-5 with 1 being impractical and 5 being highly practical.
7. How might patients from differing races/ethnicities, geographic locations, socioeconomic status, and/or educational levels find the DBT skills helpful, or not?
8. How might patients from differing races/ethnicities, geographic locations, socioeconomic status, and/or educational levels find the DBT skills practical and/or convenient, or not?

Recruitment and Participation

1. How likely would your patients be to participate in and complete this intervention?
2. What factors would encourage their recruitment and participation?
3. What are barriers to recruitment and participation?
4. What recruiting methods for support programs have been most successful with your patients?
5. What are your thoughts on other oncology clinicians and staff you know being willing to encourage patients to sign up for and participate in this intervention?
6. What additional thoughts or suggestions on recruitment and participation do you have?

General

1. How do you think our intervention could lead to smoking reduction or cessation?
2. How do you think this intervention could improve overall patient outcomes?

Thank you very much for your input on the intervention. Now I would like to move on to the patient questionnaires.

**Patient Questionnaires**

We are looking at several factors to better understand what contributes to continued and increased smoking to help inform our intervention and to measure outcomes. Some of the factors we are using for these purposes include depressive and anxious feelings, stress, the ability to tolerate and manage distress and emotions, and mental adjustment to cancer.

To assess these factors in our study, we are using these questionnaires:

**Mental Adjustment to Cancer (MAC)**

- Assesses the extent to which patients adopt the following responses to their diagnosis and treatment
  - “fighting spirit”
  - “helpless/hopeless”
  - “fatalism”
  - “avoidance”

**Difficulties in Emotion Regulation (DER)**

- asks patients how they relate to their emotions to assess for the following emotion regulation problems
  - nonacceptance of emotional responses,
  - difficulty engaging in goal-directed behavior,
  - impulse control difficulties,
  - lack of emotional awareness,
  - limited access to emotion regulation strategies
  - a lack of emotional clarity

**Distress Tolerance Scale (DTS)**

- assess the ability to withstand uncomfortable states

**Depression, Anxiety, and Stress Scale (DASS)**

- assess three related negative emotional states: depression, anxiety, and perceived stress

**Cancer Patient Tobacco Use Questionnaire**

- assesses smoking behavior and history

Questions

1. How do these factors influence your patients’ smoking behaviors?
2. In your opinion, how well do each of these assessments measure the intended factors?
3. What are some other factors not included in our assessments that you feel influence your patients’ smoking behaviors?

Other thoughts, perceptions, and/or suggestions

Lastly, based on your experiences, do you have any additional thoughts, perceptions and/or suggestions?

**Closing**

Thank you very much for meeting with me today to discuss this important topic and for contributing to our efforts to develop an effective intervention for patients.
